# Ethnic inequalities in primary care for people with multiple long-term conditions: evidence from the General Practice Patient Survey

**DOI:** 10.1101/2024.03.31.24305132

**Authors:** Brenda Hayanga, Mai Stafford, Laia Bécares

## Abstract

**Objective:** To examine the relationship between ethnicity and experiences of primary care for people with multiple long-term conditions (MLTCs) and assess the relative importance of demographic, practice, and area-level factors as influences on primary care experiences across ethnic groups.

**Design:** A retrospective study using 2018-19 GP Patient Survey data linked to the General Practice Workforce data, and the Office for National Statistics data.

**Setting:** UK

Participants: 294,987 respondents with two or more long-term conditions with complete data on selected demographic variables (age, gender, ethnicity, economic activity), practice and area-level variables (number of full-time equivalent GPs and nurses, practice size, area-deprivation, area life expectancy and ethnic density).

**Main outcome measures:** Multilevel regression analysis used to assess the relationship between ethnicity and experience of accessing primary care (i.e. satisfaction with appointment times, types and booking experience) and interacting with healthcare professionals (i.e. satisfaction with confidence and trust in healthcare professionals and the extent to which patients feel healthcare professionals listen to them, give them enough time, treat them with care and concern, involve them in healthcare decisions, and meet their needs). Separate regression models built for each outcome and included i) each covariate separately, ii) demographic factors (iii) demographic, practice, and areal-level factors.

**Results:** Upon full adjustment Arab, Bangladeshi, Chinese, Indian, Pakistani, other Asian, mixed white and Asian, other white and other ethnic group people with MLTCs have both lower levels of satisfaction with primary care access and interacting with healthcare professionals compared with white British people. The influence of demographic, practice and area-level factors is not uniform across ethnic groups. For example, demographic factors account for the inequalities in levels of satisfaction with access to primary care between white British people and Black other, mixed other, mixed white & Black Caribbean and Gypsy & Irish Travellers. However, practice and area-level factors strengthen inequalities in the experience of accessing primary care for Bangladeshi, Indian and Pakistani people.

**Conclusions:** Given that patient experience is a key aspect of healthcare quality and is said to be associated with favourable health outcomes, the inequalities identified in this study are concerning. The poorer experiences of primary care might be one mechanism by which people with MLTCs from minoritised ethnic groups have poorer health outcomes. In addition to the assessment of other practice and area-level factors, qualitative studies are required to understand and effectively address the sources of ethnic inequalities in primary care experiences for people with MLTCs.

## INTRODUCTION

Studies show that people from minoritised ethnic groups with multiple long-term conditions (MLTCs) face more disadvantage in the number, impact, and quality of care for their long-term conditions. ^1-6^ Not only do they have as many or more long-term conditions than their white counterparts but they are also more likely to receive lower continuity of care compared with white patients. ^7 1^ Further, risk of mortality for people with MLTCs is higher for minoritised ethnic groups compared with white people. ^5^ Despite this, very few studies have examined whether ethnic inequalities for people with MLTCs extend to patient experience, a key aspect of healthcare quality associated with lower readmission rates, lower mortality rates, better adherence to medication, and higher levels of trust. ^8-12^ Studies of single conditions provide compelling evidence to suggest that patients from minoritised ethnic groups report poorer experiences in primary care. ^13-17^ Yet, it is important to remember that people with MLTCs utilise healthcare services more often than people without long-term conditions.^18^ Many often juggle multiple healthcare professionals, appointments, and treatments for different health conditions concurrently, all of which is made more challenging in fragmented health and care systems which focus on single health conditions. ^19 20^

In addition to socio-demographic factors (e.g. age, gender, socioeconomic status)^17^, evidence from studies of single conditions suggest that ethnic inequalities in patient experience can also be influenced by wider factors. For example, low patient satisfaction scores among minoritised ethnic group people have been explained by practice-related factors such as low practice performance, ^16 21^ and staff and doctor’s communication skills. ^17^ Relatedly, studies have shown that general practices that serve more socio-economically deprived populations tend to have the lowest patient satisfaction^22^. Such findings are concerning because people from minoritised ethnic groups tend to be overrepresented in deprived neighbourhoods.^23^ Some studies suggest that for people from minoritised ethnic groups, decreased ethnic density is associated with increased satisfaction with health services. ^24^ However, little is known about the ways in which these wider processes impact the experiences for people with MLTCs.

Given the current emphasis on tackling healthcare inequalities and improving patient experience in the UK, ^25 26^ an investigation is required to ascertain whether there are any ethnic inequalities in patient experience for people with MLTCs. Examining how practice and area-level factors contribute to any observed inequalities is critical to understanding the modifiable, supply factors that can be addressed with policy and practice interventions to inform efforts that can improve the health of populations and reduce ethnic health inequalities for people with MLTCs. Therefore, the aims of this study are to:

1. Examine whether experiences of primary care vary across ethnic groups for people with MLTCs; and
2. Examine the relative importance of demographic, area-level, and practice-level factors as influences on primary care experiences across ethnic groups for people with MLTCs.

## METHODS

### Data

This analysis uses data from the GP Patient Survey (GPPS), the General Practice Workforce, and the Office for National Statistics (ONS). These data provide a snapshot of the primary care general practice workforce in England and include data about administrative staff, direct patient care, general practitioners, nurses, and non-clinical staff. ^27 28 29 30^ The GPPS is an independent survey conducted by Ipsos MORI on behalf of NHS (National Health Service) England. ^30^ Approximately two million patients across the UK are invited to respond to questions about their local GP services relating to awareness of, and satisfaction with services, experiences of booking appointments and care quality. ^31^ We used GPPS data from 2018 and 2019 which included respondents aged 16 years and above. ^30 31^ Key variables include age, gender, ethnicity, economic activity, long-term conditions, practice list size, practice deprivation and variables denoting patient experience of accessing and interacting/communicating with healthcare professionals. We focus on these two domains of patient experience (access and interaction/communication) based on qualitative evidence which suggests that many people with MLTCs face challenges when booking appointments for their different health conditions especially when navigating inflexible, under-resourced healthcare systems. ^32 33^ Additionally, many patients with MLTCs feel that healthcare professionals do not take enough time to explain their conditions or treatment, leaving them unable to fully understand their diagnosis, treatment, medication, or expectations of them in terms of managing their conditions. ^32 33^

We used the General Practice Workforce series of Official Statistics to extract data on the number of full time equivalent (FTE) GPs and nurses in each practice together with the practice code. We used ONS data to obtain information on ethnic group and life expectancy at the Middle Layer Super Output Area (MSOA) from the Nomis website where the ONS publishes statistics on the population, society, and the labour market at national, regional, and local levels.^34^

### Data linkage

We combined the 2018 and 2019 GP Patient Survey and linked them with the 2018/2019 General Practice Workforce data using the unique practice codes available in both datasets. Using the practice postcode, we combined this dataset with the ONS postcode directory to obtain the MSOA codes. These codes subsequently allowed linkage to area-level deprivation, area-life expectancy, and ethnic density.

### Measures

#### Patient experience

We used the responses to the questions about satisfaction with appointment times (Q25), satisfaction with appointment type (Q104) and satisfaction with the experience of making an appointment overall (Q18) to provide insight into the experiences of accessing primary care. For insight into the experiences of interacting with healthcare professionals, we used the responses to the questions about the extent to which the healthcare professional was good at giving enough time (Q86a), listening (Q86b), treating patients with care and concern (Q86e) and whether patients felt involved in decisions about care and treatment (Q88), and the extent to which the patients had confidence and trust in healthcare professional (Q89) and had their needs met (Q90) during their last appointment. We created a composite score from the three questions relating to access and another composite score from the six questions relating to interaction with healthcare professionals. We recoded responses denoted as ‘no option/doesn’t apply’ as missing and excluded them from analyses. We calculated the mean for items for all respondents who answered at least two of the three access questions and four of the six interaction questions. The composite scores were linearly rescaled to a range of 0 (denoting least favourable response) to 100 (denoting the most favourable response).

#### Patient characteristics

We extracted the age-group, gender, ethnicity, and economic activity of the respondents directly from the survey responses. We grouped the respondents into five age categories (< 35, 35-44, 45-54, 55-64, 65+), two sex categories (male and female) and five economic activity categories (retired, employed, unemployed, long-term sick/disabled and other which consisted of students, family carers and those involved in other activities). In line with the England and Wales 2011 Census ethnic categories, we used 18 ethnic categories which were self-ascribed. ^35 36^ We included only patients who reported having two or more long-term physical and/or mental health conditions. We identified respondents based on their responses to a question asking whether they had long-term conditions and if so, to select from a list of 16 long-term physical and mental health conditions (Supplementary Table 1). Respondents who reported fewer than two long-term conditions were excluded.

#### Practice and area characteristics

Through linkage with General Practice Workforce data, we obtained the number of full-time equivalent GPs and nurses. From the practice list size, we created a practice-size variable comprising of five categories (<3000, 3000-5999, 6000-8999, 9000-11999, 12000+). We recoded the Index of Multiple Deprivation scores into quintiles to denote the socio-economic deprivation of the practice. ^29 37^ We used area-life expectancy to provide an indication of the areas that have greater need.^38^ We included ethnic density in our analyses as it has been negatively correlated with satisfaction with services. ^24^ However, studies also show a positive correlation between ethnic density and social cohesion. ^39^ Ethnic density may also foster the development of positive roles,^40^ facilitate increased political mobilisation and material opportunities, and encourage healthy behaviour. ^41^ We calculated ethnic density for each ethnic group as a percentage of the total number of people within each MSOA.

### Statistical analysis

We created an analytical sample which included only people with MLTCs who had complete data on demographic, practice, and area-level variables (Figure 1). The differences in demographic characteristics between people with missing ethnicity data and people with complete ethnicity data were negligible (Supplementary Table 2). People with four or more conditions (20% vs 23%) and people living in the least deprived quintile (27% vs 30%) were overrepresented among those with missing ethnicity data compared to those with complete data (Supplementary Table 2). To analyse the relationship between ethnicity and experience of accessing primary care and interacting with healthcare professionals, we used a three-level regression analysis, with MSOA as level 3, practice as level 2 and patients as level 1 to control for potential correlation of patients within each practice, and the correlation of practices within each MSOA. This approach allowed us to explore the extent of between-practice, and between-MSOA variation in responses and to avoid overstating the importance of practice-level or area-level factors as the source of variation in patient experience.^42^ We built separate regression models for each outcome and included i) each covariate separately, ii) ethnicity, age, gender, and economic activity, iii) ethnicity, age, gender, economic activity, number of full-time equivalent GPs and nurses, practice size, area-deprivation, area life expectancy and ethnic density. After running each model, we calculated the intraclass correlation coefficient to assess the percentage of total variation in patient experience attributable to practice-level and area-level factors. Additionally, we conducted a sensitivity analysis by creating separate models for participants with and without MLTCs that include a mental health condition. We used RStudio (R04.2.0) for data linkage and Stata/MP 18 for all our analyses. ^43^

**Figure 1.**
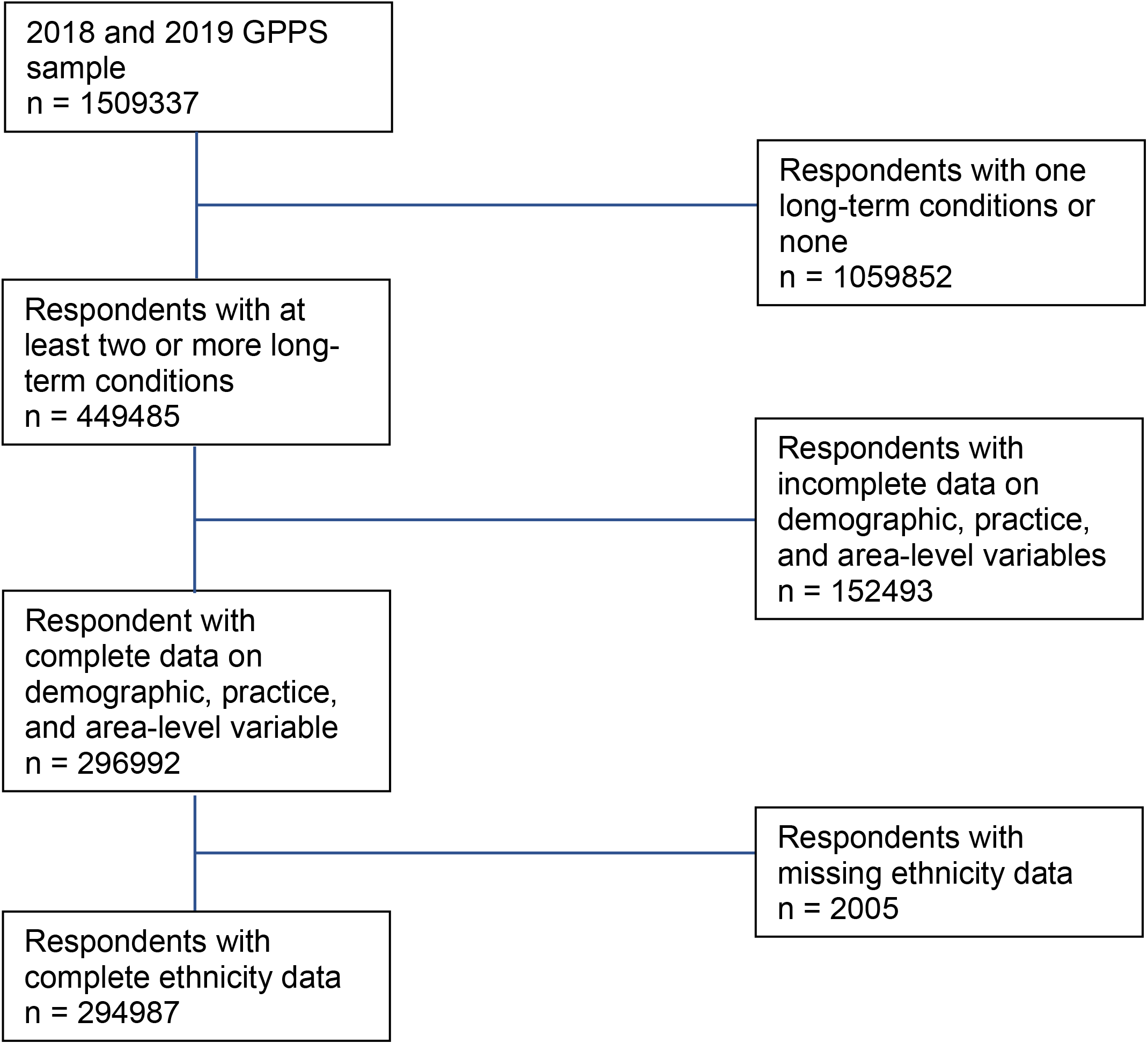
Flow chart to get analytical sample.

## RESULTS

The characteristics of the study respondents are presented in Table 1. A total of 294,987 respondents were included in the analysis, 88% of whom were of white British ethnicity. There was a higher proportion of women (52%) than men (48%) and most of the sample were aged 65 years and over (61%). Just over half the sample had two long-term conditions (53%). Only 8% of the sample were registered in practices with more than 12,000 patients. Nearly a third of the sample were registered in practices that were in the most deprived quintile (27%). The number of full-time equivalent nurses ranged from 0 to 32 with a median of 2. The number of full-time equivalent GPs ranged from 0 to 40 with a median of 4. The average area-life expectancy was 78.7 years. Ethnic density ranged from 0 to 98% with a mean of 71/%. Among minoritised ethnic groups own ethnic density ranged from 0 to 2.6% for Gypsy and Irish Travellers, to 0 to 83% for the Indian ethnic group (See supplementary Table 3).

**Table 1.** Characteristics of the study population

|  |  | **N** | **%** |
| --- | --- | --- | --- |
| **Total sample** |  | 294987 | 100 |
| **Sex** |  |  |  |
|  | Male | 142793 | 48.4 |
|  | Female | 152194 | 51.6 |
| **Age categories, years** |  |  |  |
|  | 65+ | 179634 | 60.9 |
|  | 55-64 | 63344 | 21.5 |
|  | 45-54 | 30993 | 10.5 |
|  | 35-44 | 11860 | 4.0 |
|  | 16-34 | 9156 | 3.1 |
| **Ethnicity** |  |  |  |
|  | Arab | 631 | 0.2 |
|  | Asian: Bangladeshi | 1053 | 0.4 |
|  | Asian: Chinese | 694 | 0.2 |
|  | Asian: Indian | 6764 | 2.3 |
|  | Asian: Pakistani | 3475 | 1.2 |
|  | Asian: Other Asian | 2336 | 0.8 |
|  | Black: African | 2584 | 0.9 |
|  | Black: Caribbean | 3089 | 1.1 |
|  | Black: Other | 476 | 0.2 |
|  | Mixed: White & Asian | 615 | 0.2 |
|  | Mixed: White & Black African | 290 | 0.1 |
|  | Mixed: White & Black Caribbean | 600 | 0.2 |
|  | Mixed: Other Mixed | 635 | 0.2 |
|  | Other Ethnic Group | 2431 | 0.8 |
|  | White: British | 259543 | 88.0 |
|  | White: Gypsy & Irish Traveller | 91 | 0.03 |
|  | White: Irish | 3408 | 1.2 |
|  | White: Other | 6272 | 2.1 |
| **Number of long-term conditions (LTCs)** |  |  |  |
|  | 2 LTCs | 155839 | 52.8 |
|  | 3 LTCs | 80300 | 27.2 |
|  | 4+ LTCs | 58848 | 20.0 |
| **Practice list size** |  |  |  |
|  | <3k | 44864 | 15.2 |
|  | 3k-5999 | 104409 | 35.4 |
|  | 6k-8999 | 79524 | 27.0 |
|  | 9k-11999 | 42958 | 14.6 |
|  | 12k+ | 23232 | 7.9 |
| **Area deprivation** |  |  |  |
|  | Least deprived quintile |  |  |
|  | IMD 5 | 41088 | 13.9 |
|  | IMD 4 | 48717 | 16.5 |
|  | IMD 3 | 57036 | 19.3 |
|  | IMD 2 | 67094 | 22.7 |
|  | IMD 1 | 81052 | 27.5 |
| **Continuous variables** |  | **Median** | **Range** |
|  | Number of FTE GPs | 4.1 | 0-40 |
|  | Number of FTE Nurses | 1.9 | 0-32 |
|  |  | **Mean** | **SD** |
|  | Area Life Expectancy (years) | 78.7 | 3.28 |
|  | Own Ethnic Density (%) | 70.6 | 29.5 |

*SD= standard deviation; IMD=Index of Multiple Deprivation; FTE = Full-Time Equivalent*

Satisfaction with accessing primary care and interacting with healthcare staff, rated on a scale of 0-100, for each ethnic group, are presented in Table 2. (A breakdown of the individual responses to the questions that comprise the composite score for each ethnic group is provided in Supplementary Table 3). Overall, the levels of satisfaction are higher for interacting with healthcare professionals (i.e. how healthcare providers listened, gave patients enough time, treated patients with care and concern, involved patients in healthcare decisions, met patients’ needs and were trusted by patients (86%) than for accessing primary care services (i.e. satisfaction with appointment times, types and booking experience) (80%) (Table 2). Irish people have the highest levels of satisfaction for both accessing primary care (82%) and interacting with healthcare staff (87%). Bangladeshi and Pakistani people have the lowest ratings for interacting with healthcare staff (76%) and accessing primary care services (70%).

**Table 2.**
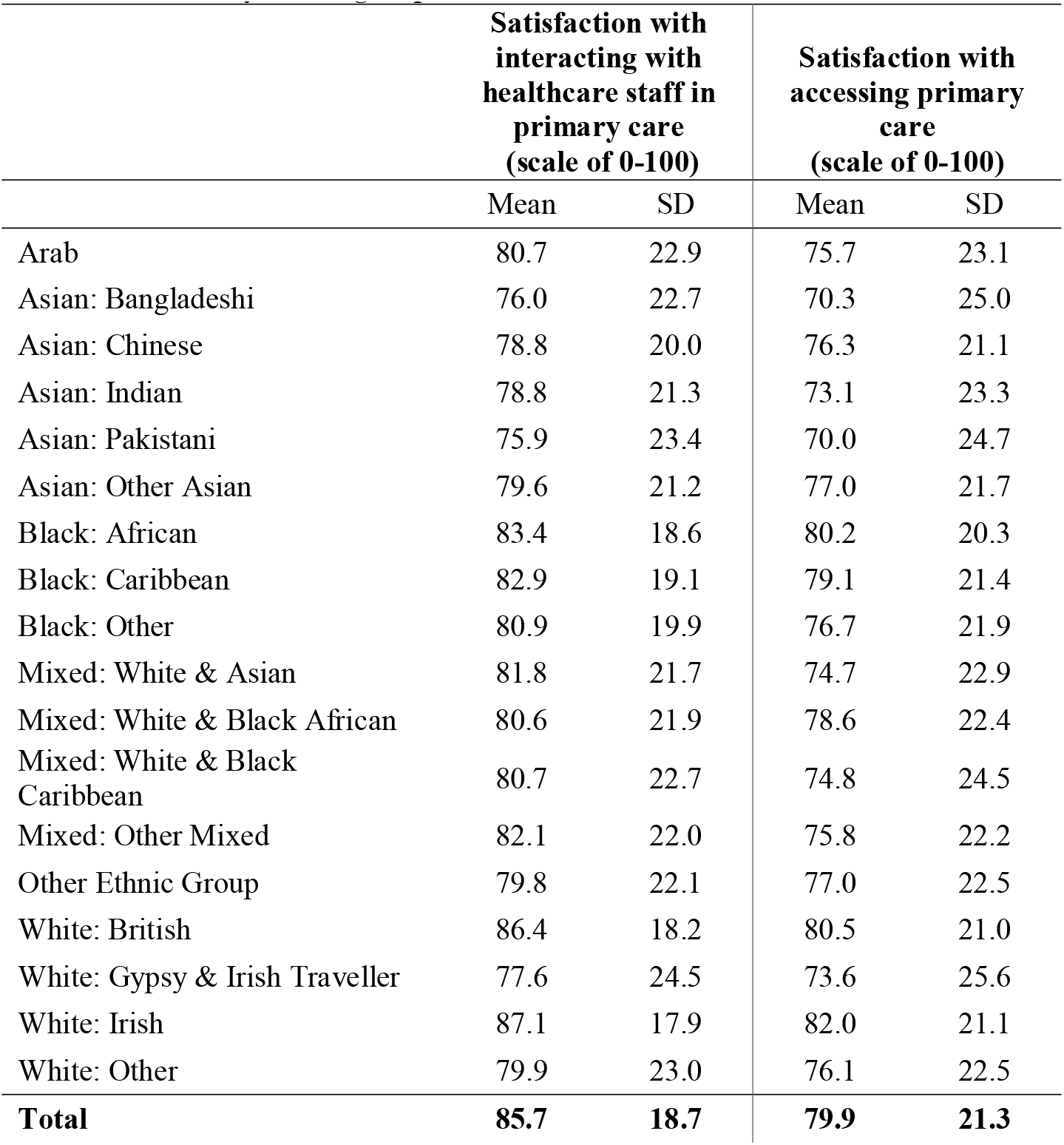
Satisfaction with accessing primary care and interacting with healthcare staff by ethnic group

|  | Satisfaction with interacting with healthcare staff in primary care (scale of 0-100) |  | Satisfaction with accessing primary care (scale of 0-100) |  |
| --- | --- | --- | --- | --- |
|  | Mean | SD | Mean | SD |
| Arab | 80.7 | 22.9 | 75.7 | 23.1 |
| Asian: Bangladeshi | 76.0 | 22.7 | 70.3 | 25.0 |
| Asian: Chinese | 78.8 | 20.0 | 76.3 | 21.1 |
| Asian: Indian | 78.8 | 21.3 | 73.1 | 23.3 |
| Asian: Pakistani | 75.9 | 23.4 | 70.0 | 24.7 |
| Asian: Other Asian | 79.6 | 21.2 | 77.0 | 21.7 |
| Black: African | 83.4 | 18.6 | 80.2 | 20.3 |
| Black: Caribbean | 82.9 | 19.1 | 79.1 | 21.4 |
| Black: Other | 80.9 | 19.9 | 76.7 | 21.9 |
| Mixed: White & Asian | 81.8 | 21.7 | 74.7 | 22.9 |
| Mixed: White & Black African | 80.6 | 21.9 | 78.6 | 22.4 |
| Mixed: White & Black Caribbean | 80.7 | 22.7 | 74.8 | 24.5 |
| Mixed: Other Mixed | 82.1 | 22.0 | 75.8 | 22.2 |
| Other Ethnic Group | 79.8 | 22.1 | 77.0 | 22.5 |
| White: British | 86.4 | 18.2 | 80.5 | 21.0 |
| White: Gypsy & Irish Traveller | 77.6 | 24.5 | 73.6 | 25.6 |
| White: Irish | 87.1 | 17.9 | 82.0 | 21.1 |
| White: Other | 79.9 | 23.0 | 76.1 | 22.5 |
| **Total** | **85.7** | **18.7** | **79.9** | **21.3** |

### Levels of satisfaction when accessing primary care services

We present the results from the regression analyses examining the association between levels of satisfaction with primary care access and socio-demographic characteristics in Table 3. Model 1 shows the unadjusted results for each covariate separately. The levels of satisfaction with the appointment times, types and booking experience are lower for women than for men.

**Table 3.**
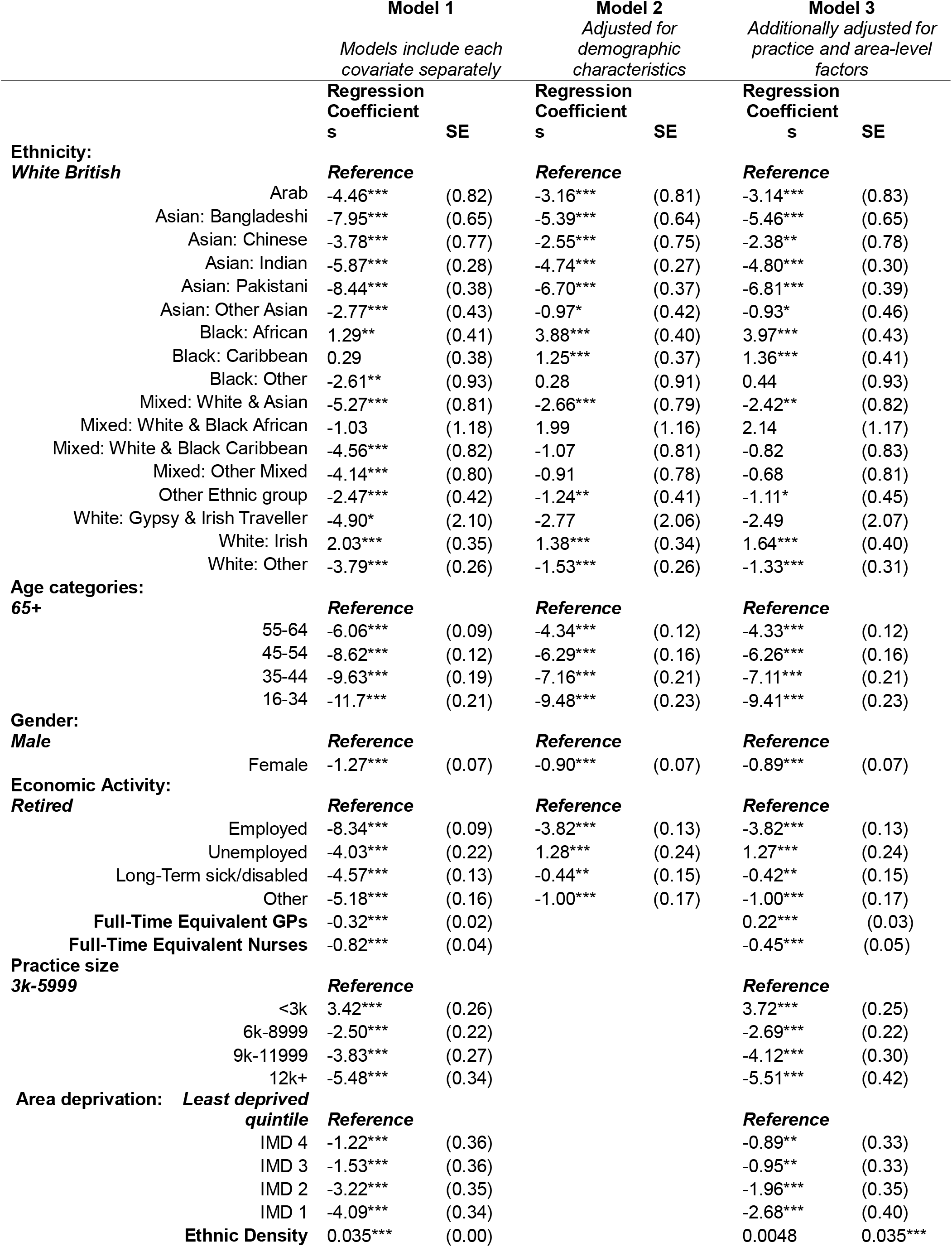
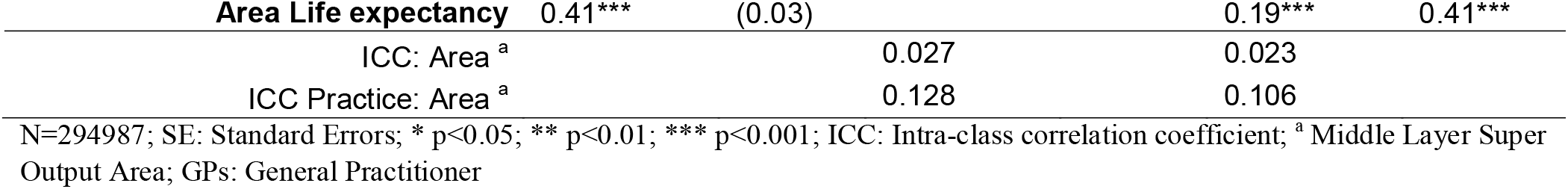
Regression analysis models showing the association between satisfaction with accessing primary care and socio-demographic characteristics

People below 65 years of age have lower levels of satisfaction when accessing primary care than people aged 65 years and above. People who are employed, unemployed, long-term sick/disabled, or engaged in other economic activities also have lower levels of satisfaction with primary care access than people who are retired. The levels of satisfaction are higher in areas of high ethnic density, and area-life expectancy but lower in practices with high area deprivation and with more full-time equivalent GPs, and nurses. Compared to practices with a list size of between 3000 and 6000 patients, those with a list size of below 3000 patients have higher levels of satisfaction with appointment times, types and booking, but those with a list size of above 6000 patients have lower levels of satisfaction (Table 3, Model 1).

When compared to white British people, all minoritised ethnic groups are less satisfied with the appointment times, types and booking experience with the exception of Black African (Regression Coefficient (Coeff): 1.29, Standard Error(SE):0.41)) and Irish people [Coeff: 2.03, SE:0.35] whose levels of satisfaction are higher, and Black Caribbean [Coeff:0.29,SE:0.38] and mixed white & Black African people [Coeff:-1.03, SE:1.18] whose levels of satisfaction with accessing primary care is not significantly different (Table 3, Model 1). Adjusting for demographic factors attenuates the effect sizes for people of Arab, Bangladeshi, Chinese, Indian, Pakistani, other Asian, mixed white & Asian, other white and other ethnicity and accounts for the differences in levels of satisfaction with primary care access for people of other Black [Coeff:0.28, SE:0.91], mixed white & Black Caribbean [Coeff:-1.07, SE:0.81], Gypsy/Irish travellers [Coeff:-2.77, SE:2.06], Other mixed ethnicity [Coeff:-0.91, SE:0.78] (Table 3, Model 2). After adjusting for demographic factors, satisfaction with accessing primary care is significantly higher for Black Caribbean people compared to white British people [Coeff:1.25, SE:0.37].

In the final model, we adjust for practice and area level factors in addition to demographic factors (Table 3, Model 3). We observe further attenuation in the strength of the association for people of Arab [Coeff: -3.14, SE:0.83], Chinese [Coeff:-2.38, SE:0.78], other Asian [Coeff: -0.93, SE:0.46], mixed white & Asian [Coeff:-2.42, SE:0.82], other white [Coeff:-1.33, SE:0.31] and other ethnicity [Coeff:-1.11, SE:0.45] but an amplification of the effect size for Bangladeshi [Coeff:-5.46, SE:0.65], Indian [Coeff:-4.80, SE:0.30], Pakistani [Coeff:-6.81, SE:0.39], all of whom report lower levels of satisfaction when accessing primary care when compared to white British people (Table 3, Model 3). In the fully adjusted model, Black African [Coeff:3.97, SE:0.43], Black Caribbean [Coeff:1.36, SE:0.41] and Irish people [Coeff:1.64, SE:0.40] report higher levels of satisfaction with primary care access than white British people (Table 3, Model 3). The intraclass coefficient in the model adjusted for all factors suggests that 10% of the total variation in levels of satisfaction with appointment types, times and booking experience is attributable to area-level and practice-level factors with the former comprising 2% of this variation (Table 3, Model 3).

An assessment of levels of satisfaction with appointment times, types and booking experience amongst people with MLTCs that include a mental health condition revealed similar patterns. After adjustment of demographic, practice and area-level factors, Bangladeshi [Coeff:-6.18, SE:1.88], Indian [Coeff:-5.39, SE:0.97] and Pakistani [Coeff:-6.77, SE:1.07] people have lower levels of satisfaction with primary care access than white British people while Black African [Coeff:5.78, SE:1.42], mixed white & Black African [Coeff:5.62, SE:2.42] and Irish people [Coeff:3.44, SE:1.12] report higher levels of satisfaction (Supplementary file 5).

### Levels of satisfaction when interacting with healthcare professionals in primary care

The results from the regression analyses examining the association between levels of satisfaction when interacting with healthcare professionals and socio-demographic characteristics are presented in Table 4. In Model 1, where each covariate was analysed separately, we notice a similar pattern to the levels of satisfaction when accessing primary care. Compared to men, women have lower levels of satisfaction with the extent to which healthcare professionals listen to patients, give them enough time, treat them with care and concern, involve them in healthcare decisions, met their needs and are considered trustworthy and confident. People who are below 65 years of age have lower levels of satisfaction than those who are 65 years and older. Practices with a list size of 12,000 patients and above have lower levels of satisfaction than practices with between 3000-6000 patients. The levels of satisfaction when interacting with healthcare professionals are lower in practices with more full-time equivalent nurses and the most deprived areas. In contrast, levels of satisfaction are higher in practices with more full-time equivalent GPs, and areas with higher ethnic density, and area life expectancy (Table 4, Model 1).

**Table 4.** Regression analysis models showing the association between satisfaction with healthcare professional interaction and socio-demographic characteristics

|  |  | **Model 1** |  | **Model 2** |  | **Model 3** |  |
| --- | --- | --- | --- | --- | --- | --- | --- |
|  |  | *Models include each covariate separately* |  | *Adjusted for demographic characteristics* |  | *Additionally adjusted for practice and area-level factors* |  |
|  |  | **Regression Coefficient** | **SE** | **Regression Coefficient** | **SE** | **Regression Coefficient** | **SE** |
|  |  | **s** |  | **s** |  | **s** |  |
| **Ethnicity:** |  |  |  |  |  |  |  |
| **White British** |  | **Reference** |  | **Reference** |  | **Reference** |  |
|  | Arab | -4.80*** | (0.74) | -3.56*** | (0.74) | -3.14*** | (0.76) |
|  | Asian: Bangladeshi | -8.64*** | (0.58) | -6.70*** | (0.58) | -6.29*** | (0.59) |
|  | Asian: Chinese | -7.17*** | (0.70) | -6.51*** | (0.69) | -5.99*** | (0.71) |
|  | Asian: Indian | -5.97*** | (0.25) | -5.40*** | (0.24) | -5.09*** | (0.27) |
|  | Asian: Pakistani | -8.59*** | (0.34) | -7.20*** | (0.34) | -6.80*** | (0.35) |
|  | Asian: Other Asian | -5.87*** | (0.39) | -4.74*** | (0.39) | -4.34*** | (0.42) |
|  | Black: African | -1.27*** | (0.37) | 0.24 | (0.37) | 0.69 | (0.39) |
|  | Black: Caribbean | -2.00*** | (0.34) | -1.50*** | (0.34) | -1.00** | (0.37) |
|  | Black: Other | -4.07*** | (0.84) | -2.36** | (0.84) | -1.83* | (0.85) |
|  | Mixed: White & Asian | -4.20*** | (0.74) | -2.43*** | (0.73) | -1.91* | (0.76) |
|  | Mixed: White & Black African | -4.93*** | (1.08) | -3.02** | (1.07) | -2.50* | (1.08) |
|  | Mixed: White & Black Caribbean | -4.73*** | (0.75) | -2.01** | (0.74) | -1.44 | (0.76) |
|  | Mixed: Other Mixed | -3.81*** | (0.73) | -1.57* | (0.72) | -1.05 | (0.74) |
|  | Other Ethnic group | -5.61*** | (0.38) | -4.66*** | (0.38) | -4.19*** | (0.41) |
|  | White: Gypsy & Irish Traveller | -7.54*** | (1.92) | -5.36** | (1.90) | -4.74* | (1.91) |
|  | White: Irish | 1.14*** | (0.32) | 0.74* | (0.32) | 1.29*** | (0.36) |
|  | White: Other | -6.10*** | (0.24) | -4.69*** | (0.24) | -4.24*** | (0.28) |
| **Age categories:** |  |  |  |  |  |  |  |
| **65+** |  | **Reference** |  | **Reference** |  | **Reference** |  |
|  | 55-64 | -2.91*** | (0.08) | -1.91*** | (0.11) | -1.88*** | (0.11) |
|  | 45-54 | -4.96*** | (0.11) | -3.58*** | (0.14) | -3.54*** | (0.14) |
|  | 35-44 | -7.19*** | (0.17) | -5.51*** | (0.20) | -5.45*** | (0.20) |
|  | 16-34 | -9.90*** | (0.20) | -8.42*** | (0.22) | -8.36*** | (0.22) |
| **Gender:** |  |  |  |  |  |  |  |
| **Male** |  | **Reference** |  | **Reference** |  | **Reference** |  |
|  | Female | -0.83*** | (0.07) | -0.49*** | (0.07) | -0.49*** | (0.07) |
| **Economic Activity:** |  |  |  |  |  |  |  |
| **Retired** |  | **Reference** |  | **Reference** |  | **Reference** |  |
|  | Employed | -4.12*** | (0.08) | -1.20*** | (0.12) | -1.22*** | (0.12) |
|  | Unemployed | -5.07*** | (0.20) | -1.36*** | (0.22) | -1.29*** | (0.22) |
|  | Long-Term sick/disabled | -3.77*** | (0.12) | -1.25*** | (0.14) | -1.17*** | (0.14) |
|  | Other | -3.92*** | (0.14) | -0.90*** | (0.16) | -0.91*** | (0.16) |
|  | **Full-Time Equivalent GPs** | 0.11*** | (0.02) |  |  | 0.32*** | (0.02) |
|  | **Full-Time Equivalent Nurses** | -0.11*** | (0.03) |  |  | -0.23*** | (0.03) |
| **Practice size** |  |  |  |  |  |  |  |
| **3k-5999** |  | **Reference** |  |  |  | **Reference** |  |
|  | <3k | 0.10 | (0.18) |  |  | 0.67*** | (0.17) |
|  | 6k-8999 | -0.12 | (0.15) |  |  | -0.79*** | (0.15) |
|  | 9k-11999 | -0.35 | (0.19) |  |  | -1.60*** | (0.21) |
|  | 12k+ | -0.92*** | (0.24) |  |  | -2.65*** | (0.30) |
| **Area deprivation:** |  |  |  |  |  |  |  |
| **Least deprived quintile** |  | **Reference** |  |  |  | **Reference** |  |
|  | IMD 4 | -0.67** | (0.23) |  |  | -0.31 | (0.21) |
|  | IMD 3 | -1.31*** | (0.23) |  |  | -0.60** | (0.21) |
|  | IMD 2 | -2.78*** | (0.22) |  |  | -1.39*** | (0.22) |
|  | IMD 1 | -3.73*** | (0.22) |  |  | -1.83*** | (0.26) |
|  | **Ethnic Density** | 0.054*** | (0.00) |  |  | 0.0083** | (0.00) |

| Area Life expectancy | 0.34*** | (0.02) |  |  | 0.12*** | (0.03) |
| --- | --- | --- | --- | --- | --- | --- |
| ICC: Area<sup>a</sup> |  |  | 0.018 | (0.01) | 0.013 | (0.01) |
| ICC Practice: Area<sup>a</sup> |  |  | 0.050 | (0.01) | 0.043 | (0.01) |

N=294987; \* p<0.05; \*\* p<0.01; \*\*\* p<0.001; ICC: Intra-class correlation coefficient; <sup>a</sup> Middle Layer Super Output; GPs: General Practitioner

When compared to their white British people, all minoritised ethnic groups have lower levels of satisfaction when interacting with healthcare professionals. Only Irish people have higher levels of satisfaction than white British people (Table 4, Model 1). After adjusting for demographic factors, we observe an attenuation of effect size for all ethnic groups and the differences in levels of satisfaction between Black African [Coeff: -0.24, SE:0.37] and white British people are no longer significant (Table 4, Model 2). After additional adjustment for area and practice level factors, we see further attenuation in the effects sizes and the levels of satisfaction with healthcare professional interaction remain significantly lower for people of Arab [Coeff:-3.14, SE:0.76], Bangladeshi[Coeff:-6.29, SE:0.59], Chinese[Coeff:-5.99, SE:0.71], Indian [Coeff:-5.09, SE:0.27], Pakistani [Coeff:-6.80, SE:0.35], other Asian [Coeff:-4.34, SE:0.42], Black Caribbean [Coeff:-1.00, SE:0.37], Black other [Coeff:-1.83, SE:0.85], mixed white & Black African [Coeff:-2.50, SE:1.08], mixed white & Asian [Coeff:-1.91, SE:0.76], Gypsy & Irish Traveller [Coeff:-4.74, SE:1.91], white other [Coeff:-4.24, SE:0.28] and other ethnicity [Coeff:-4.19, SE:0.41] (Table 4, Model 3). The levels of satisfaction when interacting with healthcare professionals remain significantly higher for Irish people [Coeff:1.29, SE:0.36] when compared to white British people (Table 4, Model 3). The intraclass coefficient in the model adjusted for all factors suggests that 4% of the total variation in the levels of satisfaction when interacting with healthcare professionals is attributable to area-level and practice level factors with area-level factors comprising 1% of this variation (Table 4, Model 3).

When we consider people with MLTCs that include a mental health condition, Bangladeshi [Coeff:-6.02, SE:1.78], Indian [Coeff:-5.92, SE:0.91], Pakistani [Coeff:-6.77, SE:1.00], other Asian [Coeff:-4.30, SE:1.31] and other white people [Coeff:-2.93, SE:0.72] have lower levels of satisfaction with healthcare professional interaction than white British people (Supplementary Table 6).

## DISCUSSION

### Principal findings

We set out to examine whether experiences of primary care vary across ethnic groups for people with MLTCs, and to understand the importance of demographic, area-level, and practice-level factors in influencing primary care experiences. To our knowledge, this is the first paper to document ethnic inequalities among people with MLTCs in the experience of accessing primary care services (i.e. satisfaction with appointment times, types and booking experience) and interacting with healthcare professionals (i.e. the extent to which patients had trust and confidence in healthcare professionals and whether they felt that they were listened to, given enough time, treated with care and concern, involved in healthcare decisions, and had their needs met). We find that the levels of satisfaction with primary care access are lower for all ethnic groups when compared to white British people, except for Black African and Irish people who have higher levels of satisfaction. Black Caribbean and mixed white & Black African people have similar levels of satisfaction to white British people. Levels of satisfaction when interacting with healthcare professionals are lower for all ethnic groups compared to white British people apart from Irish people who have higher levels of satisfaction. Studies have shown that general practices that serve more socio-economically deprived patient populations, who generally have poorer health, complex needs, and greater need for primary care, receive less funding per weighted registered patients, have difficulties with recruitment and have increased workload than practices serving more affluent patient populations.^22^They also tend to have the lowest patient satisfaction and Quality and Outcomes Framework scores and have a higher likelihood of being rated as ‘inadequate’ or ‘requires improvement’ by the Care Quality Commission. ^22^ People from minoritised ethnic groups are overrepresented in the most deprived neighbourhoods.^23^ Therefore, they are most likely to be served by poor performing practices with inadequate healthcare provision, all of which impact on their experiences.

After accounting for demographic, practice and area-level factors, people of Arab, Bangladeshi, Chinese, Indian, Pakistani, other Asian, mixed white & Asian, other white and other ethnicity not only report lower levels of satisfaction with appointment times, types and booking experience than white British people, but they also report lower levels of satisfaction with the extent to which healthcare staff listen to them, give them enough time, treat them with care and concern, involve them in healthcare decisions, meet their needs and are regarded with trust and confidence. A similar trend is observed for people of Bangladeshi, Indian and Pakistani ethnicity with MLTCs that include a mental health condition. The influence of demographic, practice and area-level factors is not uniform across ethnic groups. For example, among Black African people, the lower levels of satisfaction with healthcare staff interaction when compared to white British people are accounted for by age, gender, and economic activity. These demographic factors also account for the differences in levels of satisfaction with access to primary care between white British people and Black other, mixed other, mixed white & Black Caribbean and Gypsy & Irish Travellers. Of note is that practice and area-level factors strengthen inequalities in the experience of accessing primary care for Bangladeshi, Indian and Pakistani people. The fact that ethnic inequalities in the experience of accessing primary care and interacting with healthcare professionals remain after adjusting for demographic, practice and area-level factors suggests that these inequalities are likely to be driven by other factors.

### Study meaning

Our findings mirror those reported by others who have examined the experiences of primary care for different ethnic groups in the UK.^14 44^ These studies also find that Black African people are most likely out of all ethnic groups to have a positive experience of making a GP appointment while Asian people are least likely to have a positive experience. Similar findings are reported by Lyratzopoulos, et al. ^45^ who found that South Asian and Chinese people have less positive primary care experiences. It is important to remember that the focus of these studies was not on people with MLTCs. As such, our analyses contribute to the literature in this area by illuminating the experiences of people with MLTCs. Our finding that people with MLTCS from all Asian ethnic groups have lower levels of satisfaction with primary care access and interaction with healthcare staff is concerning. They are partially supported by Mead and Roland who examined why evaluations of primary care among minoritised ethnic groups were poorer than those of white people. ^17^ They found that people of Asian ethnicity had lower evaluations of primary care, except for Chinese patients whose differences were accounted for by issues relating to communication with practice staff. ^17^ The authors propose that the lower levels of satisfaction for Asian respondents might be the result of a higher expectation of accessing primary care. ^17^ Others suggest that the experiences of Asian patients may be driven by a lower quality of communication.^21^ Support for this notion is provided by Ahmed, et al. ^15^ whose analyses suggests that patient experience among Bangladeshi and Pakistani patients is improved where practices offer a language that is concordant with the patient’s ethnicity.

Despite language and communication issues being proffered as the reasons behind ethnic inequalities in primary care experiences for people of Asian ethnicity, these narratives dismiss the fundamental role of racism and discrimination in shaping their experiences and those of other minoritised ethnic groups.^46^ Patillo and colleagues’ review of studies examining racism and its impact on access to healthcare in Europe documents how people from minoritised ethnic groups frequently face general anti-migration bias, othering, racist language and behaviour, differential treatment, and health-related prejudices often specific to their ethnic or national category.^47^ They surmise that prejudices and stereotypes rooted in racism and coloniality can manifest in the behaviours of healthcare professionals, which in turn, translate into poor patient experience.^47^ The review highlights that studies examining the impact of racism on the primary care experiences in Europe are not only scarce, but also, the few studies that exists often focus on interpersonal racism. Given that racism also operates at a structural and institutional level,^48^ our understanding of the primary care experiences of minoritised ethnic groups (with and without MLTCs) is incomplete. Further research is warranted to address this gap to understand the role that racism, at all levels, plays in shaping the experiences of primary care.

In this study, we found that as the number of FTE nurses increased, the level of satisfaction with appointment times, types and booking experience decreased. We also found that the levels of satisfaction when interacting with healthcare staff decreased with an increase in the number of nurses but increased with an increase in the number of FTE GPs. This finding is intriguing and might reflect that patients with MLTCs might prefer to see a GP rather than a nurse for their healthcare needs. Such findings have serious implications for strategies to increase the multidisciplinary workforce aimed at reducing the workload of GPs which has increased owing to the growing number of patients with complex needs (including MLTCs), challenges with recruitment, and underinvestment in general practice. ^49^ Findings from qualitative research exploring the ways in which general practices in England operate with a work force that has a diversity of skill-mix suggest that patients do not always feel comfortable about disclosing details of their ailments to non-medical staff due to concerns about confidentiality, and/or confidence in their abilities.^50^ In light of our findings, some proposals (e.g. the use of practice nurses to undertake most routine chronic disease management or delegation of some work done by GPs to non-GP practitioners) will need careful consideration given the direct impact they may have on the growing number of people with MLTCs who utilise healthcare services more than those without MLTCs.^51 52^

With regards to area-level factors, we found that the levels of satisfaction with primary care access and interacting with healthcare professionals are better in areas of high ethnic density and areas of high life expectancy. This finding is in contrast to that of Bécares and Das-Munshi who examined the association between ethnic density and rates of health seeking behaviour, expected discrimination and satisfaction with services.^24^ Their findings suggest that decreased ethnic density is associated with increased satisfaction with health services among people from minoritised ethnic groups.^24^ Given that areas with higher ethnic density are also more deprived and health services might be poorer, the authors attribute this finding to residual confounding effects of socioeconomic indicators.^24^ The differences in findings between our study and theirs might be because our study specifically focused on satisfaction with primary care access and interaction with healthcare professionals whilst the aforementioned study measured satisfaction with local health services in general.

One way in which ethnic density impacts on healthcare experiences for minoritised ethnic groups in our study could be through the ethnic make-up of the staff in general practice. Arguably, areas of high ethnic density may have healthcare staff whose ethnic identities reflect the patient population. Some international studies suggest that racial concordance contributes to more effective therapeutic relationships and improved healthcare.^53^ Such findings underscore the importance of diversifying the ethnicity of healthcare worker.^53^ However, others find that some patients prefer to be seen by practitioners from other ethnic groups.^54^ Evidently, the effect of racial/ethnic concordance on patient experience and outcomes is complex. Further research is required to understand the mechanisms by which ethnic density influences patient experiences through ethnic dis/concordance.

### Strengths and weaknesses

This study provides a novel contribution to our understanding of ethnic inequalities in the experiences of accessing primary care services and interacting with healthcare staff for people with MLTCs. Our analyses provide strong evidence of ethnic inequalities in the experiences of primary care for people with MLTCs in domains that are important to them based on findings from qualitative studies.^32 33^ The use of 18 ethnic group categories has allowed for the identification of ethnic groups at risk of poor experiences, particularly Asian ethnic groups who have lower ratings of both accessing primary care and interacting with healthcare staff. Our findings illuminate differences between ethnic groups that are often aggregated, thereby, masking differences between ethnic groups.^3^ For example, we have shown that the experiences of accessing primary care and interacting with healthcare professionals for Black African, Black Caribbean and Black Other are not the same, yet they are often grouped together in analyses.^17^ Relatedly, the study illuminates primary care experiences of groups such as Arab people or Gypsy/Irish Travellers who are often excluded from analyses or aggregated with other people of white ethnicity despite their poor health outcomes.^4^

Our study is not without limitations. First, nearly two thirds of the sample (61%) consisted of people aged 65 years and above. Therefore, it could be argued that the experiences of primary care services reported in this study may be driven by the experiences of older people. However, our study focuses on people with MLTCs. Given that the number of long-term conditions increases with age, ^55^ a sample that consists of older people can be expected. Nevertheless, we sought to isolate the true relationship between ethnicity and experiences of primary care and remove age-related influences by controlling for age (and other factors e.g. gender, economic activity), in our analysis. Second, we constructed composite scores by combining the responses of individual measures of patient access and interaction into a single score. Concerns about the use of composite scores in statistical analyses include the lack of transparency in selecting individual measures, the likelihood of obscuring underlying measures, the failure to present uncertainly, and banding to get measures onto consistent scales^56^. However, the use of composite scores is commonplace in studies exploring patient experience ^8 15 57^. In line with these studies, we constructed composite scores and have outlined the process by which we selected the individual measures to provide clarity and transparency.

Third, analysis of how patient experiences vary by age and gender for different ethnic groups was beyond the scope of this study. Findings from analyses conducted by Burt, et al. ^58^ identified stark differences in experiences with GP-patient communication among older, female, Asian patients and younger ‘patients of other white ethnicity. Such analyses are crucial for identifying vulnerable populations who can then be targeted for further investigations to understand the reasons underlying their poor experiences. Findings from such analyses can contribute to tailored efforts to ensure they receive equitable quality care. Such studies could also examine how other practice-level factors (e.g. skills-mix) influence experiences of primary care for people with MLTCs from minoritised ethnic groups.

Fourth, in sensitivity analyses we examined ethnic inequalities in experiences of access to primary care and interaction with healthcare staff for people with and without MLTCs that include a mental health condition. We found similar patterns for those with and without an MLTCs that includes a mental health condition. However, it is important to note that when respondents of the GP Patient Survey are asked about the presence of a long-term mental health condition, they are not provided with a broad range of mental health conditions to choose from. It is possible that we may have arrived at different results were the participants asked to select a mental health condition from a wide range of mental health conditions as recorded in primary care. Future research could investigate ethnic inequalities of primary care experiences for people with MLTCs that include a mental health condition using sources of data that provide a comprehensive definition of mental health conditions.

## CONCLUSION

We have identified ethnic inequalities in the experiences of primary care services for people with MLTCs. Compared to white British people with MLTCs, Arab, Bangladeshi, Chinese, Indian, Pakistani, other Asian, mixed white & Asian, other white and other ethnic group people have both lower levels of satisfaction with appointment times, types and booking experience and lower levels of satisfaction with confidence and trust in healthcare professionals and the extent to which they feel healthcare professionals listen to them, give them enough time, treat them with care and concern, involve them in healthcare decisions, and meet their needs. These inequalities are concerning given that patient experience is a key aspect of the healthcare quality and is said to be associated with favourable health outcomes such as lower readmission rates, lower mortality rates, better adherence to medication, and higher levels of trust. ^8-12^ The poorer experiences of primary care might be one mechanism by which people with MLTCs from minoritised ethnic groups have poorer healthcare outcomes.^1-5 46^ We found that the influence of demographic, practice and area-level factors is not uniform for the different minoritised ethnic groups. This finding alerts us to the heterogeneity of minoritised ethnic groups whose experiences are also varied. It underscores the importance of adopting an intersectionality approach to understanding the reasons underlying ethnic inequalities in the experiences of primary care and the need to move away from blanket approaches to improve healthcare experiences which ignore the nuances between different minoritised ethnic groups. In addition to assessing the influence of other practice and area-level factors, qualitative studies are crucial for the identification, understanding, and formulation of solutions which will effectively address the sources of ethnic inequalities in primary care experiences for people with MLTCs from minoritised ethnic groups.

## Data Availability

The study uses individual-level data from General Practice Patient Survey (GPPS) which is available from Ipsos MORI via a data sharing agreement with NHS England. Although these data are anonymised, they are considered sensitive data in the UK by the Data Protection Act and, therefore, cannot be shared publicly. Information about applying to use data from GPPS can be found at https://gp-patient.co.uk/contact. The GPPS data was linked to data from the General Practice Workforce, and the Office for National Statistics (ONS) which is publicly available from https://digital.nhs.uk/data-and-information/publications/statistical/general-and-personal-medical-services and https://www.nomisweb.co.uk/respectively.

https://www.nomisweb.co.uk/

## DECLARATIONS

### Contributors

MS, LB and BH conceptualised the study, devised the primary research questions and analysis plan. BH conducted a review of the literature, formal statistical analysis and led the preparation of the manuscript. Output from all analyses was shared with all authors. MS, and LB critically reviewed, commented and edited the initial and subsequent manuscripts, providing methodological and intellectual feedback. BH revised subsequent drafts and submitted the manuscript for publication. All authors have read and agreed to the final version of the submitted manuscript. BH is the guarantor and attests that all listed authors meet authorship criteria and that no others meeting the criteria have been omitted.

### Ethics approval

Not applicable.

### Transparency

BH, the manuscript’s guarantor affirms that the manuscript is an honest, accurate, and transparent account of the study, that no important aspects of the study have been omitted, and that any discrepancies from the study as originally planned have been explained.

### Copyright/license for publication

The Corresponding Author has the right to grant on behalf of all authors and does grant on behalf of all authors, a worldwide licence to the Publishers and its licensees in perpetuity, in all forms, formats and media (whether known now or created in the future), to i) publish, reproduce, distribute, display and store the Contribution, ii) translate the Contribution into other languages, create adaptations, reprints, include within collections and create summaries, extracts and/or, abstracts of the Contribution, iii) create any other derivative work(s) based on the Contribution, iv) to exploit all subsidiary rights in the Contribution, v) the inclusion of electronic links from the Contribution to third party material where-ever it may be located; and, vi) licence any third party to do any or all of the above.

### Patient and public involvement

This paper is part of a series of papers from a three-year project funded by the Health Foundation from 2020 to 2023. At the start of the project, the team shared the aims and objectives of the project with the Health Foundation Inclusion Panel and sought their feedback on the ongoing reviews and the data analysis plan. They suggested that when conducting our analyses, we should consider the complexity and diversity of the patient population, wider determinants of health which shape health outcomes, the use of finer ethnic group categories where possible (because inequalities can be masked when broad categories are used) and whether inequalities are driven by a particular condition or combinations of conditions. In response, we used data from the GP Patient survey which allowed for the disaggregation of ethnic groups into 18 categories and identified patterns across these groups including Arab and Gypsy, Roma and Irish Travellers who are often excluded or included as part of broader ethnic categories. In our analyses, we considered the role of socio-demographic, practice and areal level factors as explanatory factors for ethnic inequalities. Further, when interpreting our findings, we considered the underlying mechanisms that can result in the observed ethnic inequalities, including racism and discrimination.

