## Supplementary Tables for "Ethnic inequalities in primary care for people with multiple long-term conditions: evidence from the General Practice Patient Survey"

| Supplementary Table 1: List of long-term conditions |
| --- |
| ▪ Alzheimer’s disease or other cause of dementia<br>▪ Arthritis or ongoing problem with back or joints<br>▪ Autism or autism spectrum condition<br>▪ Blindness or partial sight<br>▪ A breathing condition, such as asthma or COPD<br>▪ Cancer (diagnosis or treatment in the last 5 years)<br>▪ Deafness or hearing loss<br>▪ Diabetes<br>▪ A heart condition, such as angina or atrial fibrillation<br>▪ High blood pressure<br>▪ Kidney or liver disease<br>▪ A learning disability<br>▪ A mental health condition<br>▪ A neurological condition, such as epilepsy<br>▪ A stroke (which affects your day-to-day life)<br>▪ Another long-term condition or disability |

**Supplementary Table 2:** Participants with complete versus missing ethnicity data

| | Complete ethnicity data | Missing ethnicity data |
|---|---|---|
| N | 294987 | 2005 |
| **Gender %** | | |
| Male | 48.41 | 48.88 |
| Female | 51.59 | 51.12 |
| **Age Categories %** | | |
| 65+ | 60.9 | 62.84 |
| 55-64 | 21.47 | 21.55 |
| 45-54 | 10.51 | 11.12 |
| 35-44 | 4.02 | 2.69 |
| 16-34 | 3.1 | 1.8 |
| **Number of long-term conditions %** | | |
| 2LTCs | 52.83 | 49.63 |
| 3LTCs | 27.22 | 27.48 |
| 4+LTCs | 19.95 | 22.89 |
| **Practice size** | | |
| <3k | 15.21 | 16.61 |
| 3k-5999 | 35.39 | 37.81 |
| 6k-8999 | 26.96 | 25.29 |
| 9k-11999 | 14.56 | 12.72 |
| 12k+ | 7.88 | 7.58 |
| **Area-deprivation %** | | |
| IMD_5: Least deprived | 13.93 | 12.67 |
| IMD_4 | 16.51 | 16.71 |
| IMD_3 | 19.34 | 17.96 |
| IMD_2 | 22.74 | 22.34 |
| IMD_1 | 27.48 | 30.32 |
| **Primary care experiences (mean)** | | |
| Interaction with healthcare staff (scale of 0-100) | 85.71 | 83.39 |
| Accessing primary (scale of 0-100) | 79.92 | 77.57 |
| **Economic activity** | | |
| Retired | 56.38 | 56.56 |
| Employed | 24.46 | 21.35 |
| Unemployed | 3.03 | 3.89 |
| long-term sick/disabled | 10.06 | 11.77 |
| Other | 6.06 | 6.43 |

Supplementary Table 3. Demographic characteristics by ethnic group

| Ethnic Groups | N | % Female | % Aged 65 years and above | % with four or more LTCS | % Retired | % Living in the most deprived IMD quintile | % in practices with 12k+ patients | Mean no. of Nurses | SD | Mean no. of GPs | SD | Mean Ethnic Density | SD | Ethnic Density Range | Mean Area Life Expectancy | SD |
|---|---|---|---|---|---|---|---|---|---|---|---|---|---|---|---|---|
| Arab | 631 | 36.0 | 36.5 | 27.1 | 29.0 | 36.1 | 4.1 | 1.4 | 1.3 | 3.8 | 3.1 | 3.3 | 3.5 | 0 - 16.6 | 79.4 | 3.9 |
| Asian: Bangladeshi | 1053 | 43.0 | 26.7 | 20.5 | 20.9 | 53.6 | 4.0 | 1.5 | 1.5 | 4.0 | 3.2 | 12.9 | 14.0 | 0 - 74 | 78.3 | 3.4 |
| Asian: Chinese | 694 | 54.2 | 49.0 | 12.7 | 47.3 | 26.1 | 7.1 | 1.8 | 1.9 | 4.4 | 3.2 | 1.6 | 1.8 | 0 - 15.2 | 79.3 | 3.5 |
| Asian: Indian | 6764 | 47.6 | 49.9 | 21.3 | 41.5 | 31.3 | 5.1 | 1.5 | 1.5 | 3.7 | 2.9 | 18.7 | 18.5 | 0 - 83.2 | 78.7 | 3.2 |
| Asian: Pakistani | 3475 | 48.4 | 37.8 | 25.2 | 27.0 | 58.9 | 2.9 | 1.5 | 1.6 | 3.4 | 2.8 | 24.6 | 21.7 | 0 - 79.7 | 77.1 | 3.2 |
| Asian: Other Asian | 2336 | 49.5 | 41.8 | 17.9 | 32.2 | 26.9 | 5.6 | 1.5 | 1.6 | 4.0 | 3.1 | 5.1 | 4.2 | 0.1 - 24.3 | 79.5 | 3.0 |
| Black: African | 2584 | 53.3 | 29.9 | 13.3 | 23.5 | 48.7 | 4.6 | 1.5 | 1.6 | 4.0 | 3.4 | 10.2 | 7.2 | 0 - 34.2 | 78.3 | 3.0 |
| Black: Caribbean | 3089 | 62.5 | 50.2 | 21.0 | 46.7 | 46.9 | 5.0 | 1.6 | 1.7 | 4.1 | 3.4 | 5.1 | 3.9 | 0 - 18.2 | 78.4 | 3.0 |
| Black: Other | 476 | 54.4 | 23.5 | 17.0 | 21.6 | 44.8 | 2.9 | 1.6 | 1.5 | 4.1 | 3.2 | 1.9 | 1.5 | 0 - 6.6 | 78.3 | 3.0 |
| Mixed: White & Asian | 615 | 52.7 | 32.7 | 19.5 | 31.5 | 30.1 | 6.5 | 2.0 | 1.9 | 4.7 | 3.4 | 1.1 | 0.6 | 0.1 - 3.1 | 78.9 | 3.4 |
| Mixed: White & Black African | 290 | 56.2 | 24.5 | 17.6 | 20.0 | 42.8 | 7.6 | 1.8 | 1.9 | 4.5 | 3.7 | 0.8 | 0.5 | 0 - 2.8 | 78.5 | 3.2 |
| Mixed: White & Black Caribbean | 600 | 59.5 | 22.3 | 19.5 | 19.7 | 44.3 | 6.3 | 2.0 | 2.3 | 4.5 | 3.6 | 1.5 | 1.0 | 0.1 - 6.3 | 78.1 | 3.2 |
| Mixed: Other Mixed | 635 | 61.3 | 26.8 | 20.6 | 24.1 | 32.0 | 7.2 | 1.9 | 1.7 | 4.5 | 3.4 | 1.3 | 0.9 | 0.1 - 3.5 | 79.1 | 3.3 |
| Other Ethnic group | 2431 | 45.2 | 44.0 | 24.5 | 34.3 | 35.8 | 5.7 | 1.7 | 1.7 | 4.3 | 3.4 | 4.4 | 3.1 | 0 - 16.1 | 79.1 | 3.4 |
| White: British | 259543 | 51.6 | 63.2 | 19.9 | 59.1 | 26.1 | 8.2 | 2.5 | 2.2 | 5.0 | 3.6 | 78.7 | 20.2 | 2.1 - 98.4 | 78.7 | 3.3 |
| White: Gypsy & Irish Traveller | 91 | 48.4 | 28.6 | 30.8 | 26.4 | 39.6 | 4.4 | 2.7 | 2.5 | 4.7 | 3.4 | 0.3 | 0.4 | 0 - 2.6 | 78.1 | 3.2 |
| White: Irish | 3408 | 50.4 | 70.2 | 23.0 | 61.9 | 31.3 | 7.4 | 2.0 | 1.9 | 4.7 | 3.7 | 1.4 | 1.0 | 0 - 5.6 | 78.8 | 3.4 |
| White: Other | 6272 | 56.0 | 40.3 | 16.4 | 33.9 | 28.0 | 8.8 | 2.1 | 2.0 | 4.8 | 3.5 | 12.1 | 8.1 | 0.4 - 42.2 | 79.4 | 3.4 |
| **Total** | **294987** | **51.6** | **60.9** | **20.0** | **56.4** | **27.5** | **7.9** | **2.4** | **2.1** | **4.9** | **3.6** | **70.6** | **29.5** | **0 - 98.4** | **78.7** | **3.3** |

Supplementary Table 4. Responses to questions about experience of interacting with healthcare provider and accessing primary care by ethnic group

| | Variables used to create composite variable for experience of interaction with healthcare professionals (HCPs) | | | | | | Used to create composite variable for experience of accessing primary care | | |
|---|---|---|---|---|---|---|---|---|---|
| Ethnic Groups | Needs met? Yes definitely | Trust and confidence in HCP? %Yes definitely. | Felt involved? %Yes, definitely. | % reporting HCP very good at showing concern | % reporting HCP very good at listening | % reporting HCP very good at being patient | % Very satisfied with appointment times available | % Very satisfied with appointment booking experience | % Satisfied with appointment type |
| Arab | 58.2 | 67.5 | 54.5 | 57.2 | 58.5 | 53.9 | 29.0 | 69.3 | 30.6 |
| Asian: Bangladeshi | 51.4 | 59.1 | 47.3 | 40.7 | 42.6 | 36.9 | 21.8 | 63.8 | 21.7 |
| Asian: Chinese | 54.0 | 58.9 | 48.4 | 47.8 | 44.5 | 42.8 | 25.2 | 76.2 | 23.9 |
| Asian: Indian | 55.8 | 63.4 | 49.6 | 46.5 | 46.3 | 42.4 | 23.5 | 67.7 | 22.7 |
| Asian: Pakistani | 52.3 | 58.7 | 47.2 | 42.3 | 43.1 | 39.1 | 21.1 | 63.4 | 20.7 |
| Asian: Other Asian | 58.6 | 65.9 | 52.6 | 48.1 | 48.2 | 43.1 | 27.6 | 74.7 | 27.5 |
| Black: African | 61.8 | 72.4 | 56.2 | 58.9 | 58.9 | 52.1 | 35.5 | 72.4 | 39.9 |
| Black: Caribbean | 62.7 | 70.9 | 57.4 | 56.8 | 55.8 | 49.4 | 33.7 | 77.4 | 32.8 |
| Black: Other | 58.2 | 66.0 | 51.5 | 52.9 | 53.2 | 47.3 | 31.9 | 70.8 | 100.0 |
| Mixed: White & Asian | 61.6 | 68.8 | 58.5 | 56.6 | 54.3 | 52.9 | 30.4 | 69.3 | 24.6 |
| Mixed: White & Black African | 57.9 | 67.2 | 53.1 | 53.1 | 57.6 | 53.1 | 36.6 | 75.9 | 34.5 |
| Mixed: White & Black Caribbean | 58.3 | 65.7 | 56.7 | 57.8 | 57.7 | 53.7 | 33.7 | 68.7 | 29.8 |
| Mixed: Other Mixed | 60.0 | 68.7 | 61.3 | 60.6 | 59.1 | 54.8 | 31.8 | 71.3 | 26.9 |
| Other Ethnic group | 58.8 | 64.2 | 54.1 | 52.9 | 53.2 | 46.7 | 30.2 | 72.9 | 32.4 |
| White: British | 69.7 | 77.0 | 67.1 | 65.0 | 63.7 | 60.5 | 39.0 | 80.6 | 33.4 |
| White: Gypsy & Irish Traveller | 57.1 | 62.6 | 48.4 | 53.9 | 52.8 | 48.4 | 27.5 | 67.0 | 100.0 |
| White: Irish | 72.9 | 77.9 | 68.4 | 66.6 | 63.9 | 62.4 | 43.0 | 80.4 | 40.4 |
| White: Other | 59.1 | 65.6 | 56.0 | 54.4 | 53.9 | 49.0 | 30.8 | 72.7 | 26.3 |
| **Total** | **68.4** | **75.7** | **65.6** | **63.5** | **62.4** | **59.0** | **37.9** | **79.5** | **32.8** |

Supplementary Table 5: Regression models showing levels of satisfaction with access to primary care for people with and without MLTCs that includes a mental health condition. *Models adjusted for demographic, practice and area-level factors*

| | | Model includes participants with a mental health condition | | Model includes participants with physical health conditions | |
|---|---|---|---|---|---|
| | | Regression Coefficients | SE | Regression Coefficients | SE |
| Ethnicity: | *White British* | *Reference* | | *Reference* | |
| | Arab | -4.08 | (2.15) | -2.76** | (0.90) |
| | Asian: Bangladeshi | -6.18** | (1.88) | -5.37*** | (0.69) |
| | Asian: Chinese | 0.37 | (2.89) | -2.52** | (0.80) |
| | Asian: Indian | -5.39*** | (0.97) | -4.79*** | (0.31) |
| | Asian: Pakistani | -6.77*** | (1.07) | -7.07*** | (0.41) |
| | Asian: Other Asian | -0.72 | (1.38) | -0.89 | (0.48) |
| | Black: African | 5.78*** | (1.42) | 3.76*** | (0.45) |
| | Black: Caribbean | 0.90 | (1.25) | 1.49*** | (0.42) |
| | Black: Other | 3.90 | (2.29) | -0.26 | (1.02) |
| | Mixed: White & Asian | -0.72 | (1.77) | -2.48** | (0.95) |
| | Mixed: White & Black African | 5.62* | (2.42) | 1.07 | (1.38) |
| | Mixed: White & Black Caribbean | -2.01 | (1.61) | 0.36 | (1.01) |
| | Mixed: Other Mixed | 0.59 | (1.67) | -0.69 | (0.95) |
| | Other Ethnic group | -0.40 | (1.16) | -1.14* | (0.48) |
| | White: Gypsy & Irish Traveller | -6.76 | (3.70) | 1.11 | (2.65) |
| | White: Irish | 3.44** | (1.12) | 1.68*** | (0.43) |
| | White: Other | -0.78 | (0.77) | -1.16*** | (0.34) |
| Age categories: | *65+* | *Reference* | | *Reference* | |
| | 55-64 | -2.26*** | (0.41) | -4.41*** | (0.13) |
| | 45-54 | -4.11*** | (0.45) | -6.28*** | (0.18) |
| | 35-44 | -4.99*** | (0.50) | -6.89*** | (0.27) |
| | 16-34 | -7.65*** | (0.51) | -8.62*** | (0.33) |
| Gender: | *Male* | *Reference* | | *Reference* | |
| | Female | -1.14*** | (0.22) | -0.79*** | (0.08) |
| Economic Activity: | *Retired* | *Reference* | | *Reference* | |
| | Employed | -4.12*** | (0.44) | -3.78*** | (0.13) |
| | Unemployed | 1.14* | (0.55) | 1.36*** | (0.29) |
| | Long-Term sick/disabled | -0.62 | (0.43) | -0.17 | (0.17) |
| | Other | -1.22* | (0.52) | -0.93*** | (0.18) |
| | Full-Time Equivalent GPs | 0.50*** | (0.06) | 0.22*** | (0.03) |
| | Full-Time Equivalent Nurses | -0.73*** | (0.09) | -0.42*** | (0.05) |
| Practice size: | *3k-5999* | *Reference* | | | |
| | <3k | 4.32*** | (0.44) | 3.75*** | (0.26) |
| | 6k-8999 | -3.33*** | (0.38) | -2.69*** | (0.22) |
| | 9k-11999 | -5.37*** | (0.52) | -4.04*** | (0.30) |
| | 12k+ | -6.56*** | (0.76) | -5.58*** | (0.43) |
| Area deprivation: | *Least deprived quintile* | *Reference* | | | |
| | IMD 4 | -1.01 | (0.54) | -0.89** | (0.33) |
| | IMD 3 | -0.89 | (0.54) | -0.90** | (0.33) |
| | IMD 2 | -1.97*** | (0.56) | -1.87*** | (0.34) |
| | IMD 1 | -3.01*** | (0.63) | -2.46*** | (0.39) |
| | Ethnic Density | 0.018** | (0.01) | 0.0082* | (0.00) |
| | Area Life expectancy | 0.16** | (0.06) | 0.19*** | (0.04) |
| | **N** | 45584 | | 249403 | |
| | ICC: Area[a] | 0.17 | (0.004) | 0.02 | (0.002) |
| | ICC Practice: Area[a] | 0.10 | (0.004) | 0.11 | (0.002) |

Supplementary Table 6: Regression models showing levels of satisfaction with healthcare provider interaction for people with and without MLTCs that includes a mental health condition. *Models adjusted for demographic, practice and area-level factors.*

| | | Model includes participants with a mental health condition | | Model include participants with physical health conditions | |
|---|---|---|---|---|---|
| | | Regression Coefficients | SE | Regression Coefficients | SE |
| Ethnicity: | *White British* | *Reference* | | *Reference* | |
| | Arab | -3.43 | (2.04) | -3.06*** | (0.81) |
| | Asian: Bangladeshi | -6.02*** | (1.78) | -6.59*** | (0.62) |
| | Asian: Chinese | -0.77 | (2.76) | -6.47*** | (0.72) |
| | Asian: Indian | -5.92*** | (0.91) | -5.18*** | (0.28) |
| | Asian: Pakistani | -6.77*** | (1.00) | -7.10*** | (0.36) |
| | Asian: Other Asian | -4.30** | (1.31) | -4.40*** | (0.43) |
| | Black: African | 1.04 | (1.35) | 0.45 | (0.40) |
| | Black: Caribbean | -1.81 | (1.18) | -0.98** | (0.38) |
| | Black: Other | -0.71 | (2.18) | -2.09* | (0.92) |
| | Mixed: White & Asian | -0.97 | (1.68) | -1.73* | (0.86) |
| | Mixed: White & Black African | -0.93 | (2.31) | -2.68* | (1.25) |
| | Mixed: White & Black Caribbean | -0.60 | (1.53) | -1.52 | (0.92) |
| | Mixed: Other Mixed | -0.96 | (1.59) | -0.52 | (0.86) |
| | Other Ethnic group | -2.11 | (1.10) | -4.57*** | (0.43) |
| | White: Gypsy & Irish Traveller | -3.97 | (3.53) | -4.75* | (2.41) |
| | White: Irish | 1.60 | (1.06) | 1.47*** | (0.38) |
| | White: Other | -2.93*** | (0.72) | -4.38*** | (0.30) |
| Age categories: | *65+* | *Reference* | | *Reference* | |
| | 55-64 | -0.62 | (0.39) | -1.84*** | (0.12) |
| | 45-54 | -2.20*** | (0.43) | -3.39*** | (0.16) |
| | 35-44 | -4.03*** | (0.48) | -5.24*** | (0.25) |
| | 16-34 | -7.82*** | (0.49) | -7.04*** | (0.30) |
| Gender: | *Male* | *Reference* | | *Reference* | |
| | Female | 0.62** | (0.21) | -0.66*** | (0.07) |
| Economic Activity: | *Retired* | *Reference* | | *Reference* | |
| | Employed | -0.55 | (0.42) | -1.44*** | (0.12) |
| | Unemployed | -1.22* | (0.52) | -0.94*** | (0.27) |
| | Long-Term sick/disabled | -1.30** | (0.41) | -0.65*** | (0.16) |
| | Other | -1.22* | (0.49) | -0.72*** | (0.17) |
| | Full-Time Equivalent GPs | 0.60*** | (0.05) | 0.29*** | (0.02) |
| | Full-Time Equivalent Nurses | -0.38*** | (0.07) | -0.21*** | (0.04) |
| Practice size: | *3k-5999* | *Reference* | | | |
| | <3k | 0.34 | (0.37) | 0.77*** | (0.17) |
| | 6k-8999 | -1.28*** | (0.32) | -0.76*** | (0.15) |
| | 9k-11999 | -3.01*** | (0.43) | -1.43*** | (0.21) |
| | 12k+ | -4.02*** | (0.63) | -2.56*** | (0.30) |
| Area deprivation: | *Least deprived quintile* | *Reference* | | | |
| | IMD 4 | -0.039 | (0.45) | -0.35 | (0.21) |
| | IMD 3 | -0.40 | (0.45) | -0.60** | (0.21) |
| | IMD 2 | -1.01* | (0.46) | -1.39*** | (0.22) |
| | IMD 1 | -1.87*** | (0.52) | -1.67*** | (0.26) |
| | Ethnic Density | 0.023*** | (0.01) | 0.13*** | (0.03) |
| | Area Life expectancy | 0.12* | (0.05) | 78.2*** | (2.20) |
| | N | 45584 | | 249403 | |
| | ICC: Area[a] | 0.007 | (0.003) | 0.014 | (0.001) |
| | ICC Practice: Area[a] | 0.043 | (0.003) | 0.043 | (0.001) |
